# The Prophylactic Effect of Oral Olanzapine on Postoperative Nausea and Vomiting in Laparoscopic Gynecological Surgery: A Randomized Clinical Trial

**DOI:** 10.1101/2020.11.27.20239582

**Authors:** Hiroyuki Seki, Norifumi Kuratani, Toshiya Shiga, Norikazu Miura, Ichiro Kondo, Mao Kinoshita, Yoshiki Sento, Yuya Masuda, Sathoshi Ideno, Kohei Ikeda, Koji Takano, Yuuki Kuwabara, Rie Kinugasa, Fumimasa Amaya, Teiji Sawa, Kazuya Sobue, Hiroshi Morisaki

**Affiliations:** Department of Anesthesiology, Keio University School of Medicine, Tokyo, Japan; Department of Anesthesiology, Kyorin University School of Medicine, Tokyo, Japan; Department of Anesthesiology, Saitama Children’s Medical Center, Saitama Japan; Department of Anesthesiology, International University of Health and Welfare, School of Medicine, Chiba, Japan; Department of Anesthesiology, Showa University School of Medicine, Tokyo Japan; Department of Anesthesiology, Jikei University School of Medicine, Tokyo, Japan; Department of Anesthesiology, Kyoto Prefectural University of Medicine, Kyoto, Japan; Department of Anesthesiology and Intensive Care Medicine, Nagoya City University, Nagoya, Japan; Department of Pain Management and Palliative Care Medicine, Kyoto Prefectural University of Medicine, Kyoto Japan

## Abstract

**Background:** Although the antiemetic effect of olanzapine on chemotherapy-induced nausea and vomiting has been characterized, the prophylactic role of olanzapine on postoperative nausea and vomiting (PONV) has not been fully elucidated. This clinical trial aims to examine the effectiveness of olanzapine for preventing PONV.

**Methods:** Patients undergoing laparoscopic gynecological surgery at 5 university hospitals in Japan will be randomly assigned to receive either 5 mg of oral olanzapine or placebo 2 hours before the induction of anesthesia. All patients will receive intravenous dexamethasone at the induction of anesthesia. The primary outcome will be the incidence of postoperative nausea and vomiting within 24 hours after surgery. Secondary endpoints will include longitudinal changes in the incidence of postoperative nausea and vomiting and overall patient satisfaction.

**Discussion:** This trial will provide a high quality evidence whether olanzapine prevents PONV in gynecological laparoscopy patients at a high risk for PONV.

**Ethics and dissemination:** The trial was approved by the institutional review board of the each participating study site. Study findings will be disseminated through peer-reviewed publications and in conference presentations.

**Trial registration:** UMIN Clinical Trials Registry (UMIN-CTR) ID: 000022634, Registered on October 1, 2016 https://upload.umin.ac.jp/cgi-open-bin/ctr/ctr_view.cgi?recptno=R000026031

---

**Multicenter Study on the Effectiveness of Olanzapine for Preventing Postoperative Nausea and Vomiting: A Clinical Trial Protocol**

Department of Anesthesiology, Keio University School of Medicine

◯ UMIN ID: 000022634
◯ Ethics Committee Approval Number: 20160154
◯ Principal Investigator Name: Hiroyuki Seki Affiliation: Dept. of Anesthesiology, Keio University School of Medicine Position: Assistant Professor Contact information: Address: 35 Shinanomachi, Shinjuku-ku, Tokyo, 160-5852, Japan TEL: +81-3-5363-3107 E-mail address:
◯ Trial Officer Name: Yuya Masuda Affiliation: Dept. of Anesthesiology, Keio University School of Medicine Position: Assistant Professor Contact information: Address: 35 Shinanomachi, Shinjuku-ku, Tokyo, 160-5852, Japan TEL: +81-3-5363-3107 FAX: +81-3-3356-8439 E-mail address

**Dates of drafts and revisions**

➢ 5/30/2016: Initial draft completed (Ver. 1)
➢ 10/14/2016: Ver. 1.1 drafted: version update due to the partial revision of the briefing form. (Addition of a grant from Japan Society of Anesthesiology.)
➢ 12/7/2016: Ver. 1.2 drafted: Addition of a grant from MSD K.K. to section 12.5: Conflicts of interest; revision of investigator from Hiroshi Morisaki to Hiroyuki Seki.
➢ 3/1/2017: Ver. 2.1 drafted: revision of olanzapine dosage (10 mg to 5 mg); revision of timing of doses (from 4 h to 2 h prior to scheduled induction of anesthesia); addition of preoperative drowsiness to potential risks; addition of “pronounced preoperative drowsiness resulting in difficulty walking to the operating room” as an indicator in safety assessment; addition of “aged ≥65 years” and “currently nursing” to the exclusion criteria; amending the withdrawal criterion of “after providing consent, the subject is determined not to meet the eligibility criteria prior to administration of the investigational agent” with “in this event, the subject shall be substituted (additional enrollment)”; addition of addenda.
➢ 8/1/2017: Ver. 2.2 drafted: version update in accordance with a supplement to address an omission from the briefing form.
➢ 3/1/2018: Ver. 3.0 drafted: version update due to revisions of the investigator regulatory managers, collaborators, and the briefing form.

## 0. Summary

### 0.1. Objective

This clinical trial aims to examine the effectiveness of the atypical antipsychotic olanzapine for preventing postoperative nausea and vomiting (PONV).

### 0.2. Subjects

Subjects are patients aged ≥20 years undergoing laparoscopic surgery for benign gynecological diseases, scheduled to last ≥1 h at any of the study centers. They must also have an Apfel simplified risk score* of ≥2.

*Consists of four items: 1) female sex, 2) history of motion sickness or PONV, 3) no smoking for ≥1 year, 4) use of postoperative opioids.

### 0.3. Investigational therapy

Two hours before scheduled induction of anesthesia, subjects will be orally administered a double-blind test capsule containing either olanzapine (5 mg) or placebo (lactose).

### 0.4. Target sample size and study period

Target sample size: 220 patients (olanzapine group: 110 patients, placebo group: 110 patients).

Study period: Date of approval – 3/31/2019

### 0.5. Trial Officer

∘ Name: Yuya Masuda
∘ Affiliation: Dept. of Anesthesiology, Keio University School of Medicine
∘ Position: Assistant Professor
∘ Address: 35 Shinanomachi, Shinjuku-ku, Tokyo 160-5852, Japan
∘ TEL: 03-5363-3107
∘ FAX: 03-3356-8439
∘

## 1. Outline

### 1.1 Trial information

∘ Title: Multicenter Study on the Effectiveness of Olanzapine for Preventing Postoperative Nausea and Vomiting
∘ Ethics committee approval no.: 20160154
∘ Clinical trial registration number: UMIN000022634
∘ The initial draft (Ver. 1) of this protocol was written on: 5/30/2016

### 1.2 Principal investigator

∘ Name: Hiroyuki Seki
∘ Affiliation: Dept. of Anesthesiology, Keio University School of Medicine
∘ Position: Assistant Professor
∘ Address: 35 Shinanomachi, Shinjuku-ku, Tokyo 160-5852, Japan
∘ TEL: +81-3-5363-3107
∘

### l.3 Trial officer

∘ Name: Yuya Masuda
∘ Affiliation: Dept. of Anesthesiology, Keio University School of Medicine
∘ Position: Assistant Professor
∘ Address: 35 Shinanomachi, Shinjuku-ku, Tokyo 160-5852, Japan
∘ TEL: +81-3-5363-3107
∘ FAX: +81-3-3356-8439
∘

### 1.4 Medical expert

The principal investigator will make medical judgments pertaining to the overall clinical trial group.

### 1.5 Study centers, directors, and regulatory managers

Center 1

∘ Name: Dept. of Anesthesiology, Keio University School of Medicine
∘ Address: 35 Shinanomachi, Shinjuku-ku, Tokyo 160-5852, Japan
∘ TEL: +81-3-5363-3107
∘ Director: Hiroshi Morisaki
∘ Position: Professor
∘
∘ Regulatory manager: Yuya Masuda
∘ Position: Assistant Professor
∘

Center 2

∘ Name: Dept. of Anesthesiology, Showa University School of Medicine
∘ Address: 1-5-8 Hatanodai, Shinagawa-ku, Tokyo 142-8555, Japan
∘ TEL: +81-3-3784-8575
∘ Director: Hiroshi Otake
∘ Position: Professor
∘
∘ Regulatory manager: Hironobu Ueshima
∘ Position: Lecturer
∘

Center 3

∘ Name: Dept. of Anesthesiology, Jikei University School of Medicine
∘ Address: 3-25-8 Nishi-Shimbashi, Minato-ku, Tokyo 105-8461, Japan
∘ TEL: +81-3-3433-1111 (Ext. 4040)
∘ Director: Shoichi Uezono
∘ Position: Professor
∘
∘ Regulatory manager: Ichiro Kondo
∘ Position: Associate Professor
∘

Center 4

∘ Name: Dept. of Anesthesiology and Intensive Care Medicine,
∘ Nagoya City University Graduate School of Medical Sciences
∘ Address: 1 Kawasumi, Mizuho-cho, Mizuho-ku, Nagoya 467-8601, Japan
∘ TEL: +81-52-851-5511 (Ext. 2288)
∘ Director: Yoshiki Sento
∘ Position: Assistant Professor
∘
∘ Regulatory manager: Yoshiki Sento
∘ Position: Assistant Professor
∘

Center 5

∘ Name: Dept. of Anesthesiology,
∘ Kyoto Prefectural University of Medicine
∘ Address: Kajii-cho, Kawaramachi-Hirokoji, Kamigyo-ku,
∘ Kyoto 602-8566, Japan TEL: +81-75-251-5633
∘ Director: Fumimasa Amaya
∘ Position: Associate Professor
∘
∘ Regulatory manager: Fumimasa Amaya
∘ Position: Associate Professor
∘

## 2. Background information

### 2.1 Investigational product

#### Name

Generic name: Olanzapine

Trade name: Zyprexa

#### Summary

Olanzapine is an atypical antipsychotic approved as a therapeutic agent for schizophrenia in the United States and Japan in 1996 and 2001, respectively. The drug was also approved in both countries for manic depressive symptoms of bipolar disorder during 2012.

### 2.2 Non-clinical studies and previous clinical trials

Olanzapine exerts antiemetic effects through simultaneously blocking multiple receptors, such as dopamine, serotonin, histamine, α_1_-adrenergic, and muscarinic receptors^1)^. Thus, the drug is recommended for preventing chemotherapy-induced nausea and vomiting (CINV) in the 2015 edition of the globally consulted National Comprehensive Center Network Guidelines^2)^ and is used for that purpose in multiple countries (including Japan). However, olanzapine has not been approved as an antiemetic.

### 2.3 Risks and benefits to subjects

#### Potential risks

According to the package insert, negative reactions from olanzapine treatment is associated with high blood glucose, leading to fatal diabetic ketoacidosis and diabetic coma. In addition, the United States Food and Drug Administration has stated that in rare cases, olanzapine can cause serious skin symptoms known as drug-induced hypersensitivity syndrome, with eosinophilia and systemic symptoms. A non-Japanese phase III trial reported the adverse reaction of transient drowsiness after investigating olanzapine effectiveness for preventing CINV. However, other adverse reactions such as hyperglycemia and weight gain were not reported^3)^. Another trial at Keio University Hospital (“Safety of olanzapine for the prevention of postoperative nausea and vomiting,” Ethics Committee approval number 20130415) found no adverse events directly due to olanzapine. A preliminary study (“Prevention of postoperative nausea and vomiting by olanzapine,” Ethics Committee approval number 20140303) of 30 patients receiving olanzapine found dysphoria in three patients and drowsiness among one of these three. However, the trial could not determine whether olanzapine directly caused these adverse events. During the present trial, three subjects who received 10 mg olanzapine experienced pronounced preoperative drowsiness, resulting in the need for wheelchair or stretcher assistance when moving to the operating room. Besides drowsiness, other adverse effects including weight gain have been associated with olanzapine use, but only after 6 months or more^3)^. Therefore, the risk of these negative side effects are extremely low after only a single administration.

#### Benefits

Non-Japanese clinical trials have shown that olanzapine prevents CINV^4)^ and PONV^5)^. Therefore, in the present study, administering olanzapine may reduce PONV incidence and increase postoperative patient satisfaction.

#### Burden on subjects

Olanzapine or placebo must be orally administered 2 h before scheduled anesthesia induction. (If surgery begins in the morning, administration must occur earlier in the morning.) Subjects are given two surveys, the first regarding PONV presence or absence at 0.5, 2, 6, and 24 h after extubation, the second regarding nausea in the morning on the second postoperative day. Drug costs are borne by the study group and patients do not incur financial burdens.

#### Overall assessment

In this study, olanzapine is administered only once, before surgery. Therefore, the risk of hyperglycemia and other adverse events is extremely low. Despite potential preoperative drowsiness, olanzapine is highly likely to prevent PONV, meaning that participation in the study should to pose no disadvantage to patients.

### 2.4 Route and dosage of administration

Two hours prior to the scheduled start of anesthesia, subjects will be orally administered one double-blind test capsule containing either olanzapine (5 mg) or lactose as a placebo.

### 2.5 Governing rules

This study will be conducted in accordance with the Declaration of Helsinki and Ethical Guidelines for Medical and Health Research involving Human Subjects.

### 2.6 Trial subject population

Subjects are patients aged ≥20 years undergoing laparoscopic surgery scheduled to last ≥1 h for benign gynecological diseases at any of the study centers. Individuals must also have an Apfel simplified risk score* of ≥2.

*Consists of four items: 1) female sex, 2) history of motion sickness or PONV, 3) no smoking for ≥1 year, 4) use of postoperative opioids.

### 2.7 References

1. Prommer E. Am J Hosp Palliat Care 2013; 30:75-82
2. http://www.nccn.org/about/news/ebulletin/ebulletindetail.aspx?ebulletinid=6
3. Navari, R.M. Expert Opin Drug Saf 2016;15:343-56
4. Chow R, et al. Support Care Cancer 2016;24:1001-8
5. Ibrahim M, et al. Egypt J Anaesth 2013;29:89-95

## 3. Objective

### 3.1 Background

Postoperative nausea and vomiting, along with postoperative pain, is a frequent complication of anesthesia, occurring in 20−30% of most patients and as many as 70−80% in high-risk patients (Table 1)^1)^. Thus, PONV is a major factor in reduced perioperative patient satisfaction. Many academic societies recommend prophylactic administration of antiemetics based on the patient risk^2,3)^. However, monotherapy with any single agent reveal limited effects. Therefore, the use of multiple antiemetics is recommended for patients with a high PONV risk.

**Table 1:**
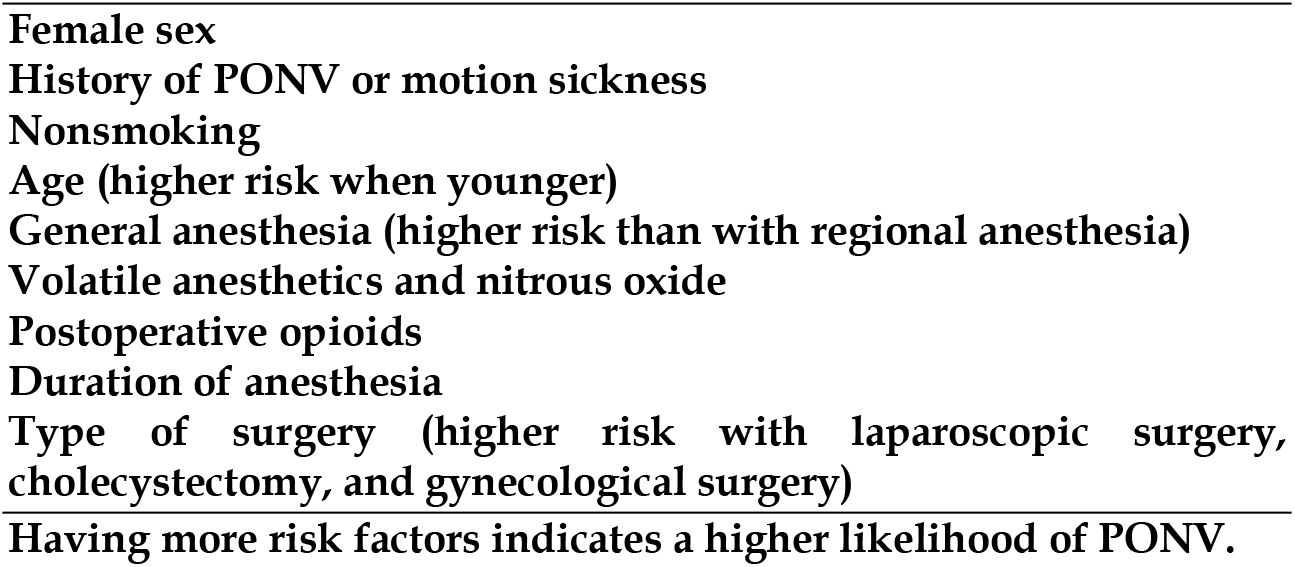
Risk factors for PONV (modified from Reference 2)

Because olanzapine has a long half-life (∼30 h) in blood^5)^, we hypothesized that a single dose could prevent PONV. However, this potential must be investigated beyond a single study^8)^. Therefore, we conducted a multicenter trial that examines olanzapine effectiveness for PONV prevention in high-risk patients.

Before conducting the present study, we conducted two preliminary trials: one to confirm olanzapine safety for perioperative patients, and another to examine the optimal timing of administration for preventing PONV.

In the trial “Safety of olanzapine for the prevention of postoperative nausea and vomiting” (UMIN000013908, Ethics Committee approval number 20130415), patients were orally administered olanzapine at concentrations of 2.5 mg (n = 10), 5.0 mg (n = 10), or 10 mg (n = 10) when leaving their ward on surgery day. We confirmed that none of the following occurred post-surgery: arrhythmia, decreased blood pressure, extrapyramidal symptoms, or abnormal drowsiness within 48 h; hyperglycemia the day after. Next, in the trial “Prevention of postoperative nausea and vomiting by olanzapine (a preliminary study)” (UMIN000016710, Ethics Committee approval number 20140313), we examined PONV incidence within 24 h post-surgery in each of the following interventions: 1) oral olanzapine (10 mg) the day before surgery (n = 10), 2) oral olanzapine (10 mg) 4 h before anesthesia (n = 10), 3) oral olanzapine (10 mg) at 9:00 PM the day before surgery and 5 mg when leaving the ward on surgery day (n = 10). Incidence of PONV in these three interventions was 5/9 (56%), 3/7 (43%), and 6/9 (67%), respectively. Notably, PONV incidence in Intervention 2 was lower than we had expected (60−80%), but overall, our findings suggested that oral administration of olanzapine 4 h before anesthesia can reduce PONV incidence.

1. Gan TJ, et al. JAMA 2002; 287:1233-1236
2. Gan TJ, et al. Anesth Analg 2014; 118:85-113
3. Apfelbaum JL, et al. Anesthesiology 2013; 118:291-307
4. Prommer E. Am J Hosp Palliat Care 2013; 30:75-82
5. http://www.info.pmda.go.jp/downfiles/ph/PDF/530471_1179044F4028_1_23.pdf
6. Tageja N, et al. Postgrad M J 2016; 92:34-40
7. Yamakawa S, Symptom management in cancer patients 2013; 24:119-125
8. Ibrahim M, et al. Egypt J Anaesth 2013; 29:89-95

### 3.2 Objective

This study aims to examine whether olanzapine prevents PONV in gynecological laparoscopy patients at a high risk for PONV.

## 4. Trial design

### 4.1 Endpoints

#### (1) Primary endpoint

The primary endpoint is the incidence of PONV within 24 h post-extubation.

#### (2) Secondary endpoints

Secondary outcomes include time to first PONV, PONV incidence, vomiting, worst nausea VAS score, significant nausea incidence (VAS score ≥40), use of rescue metoclopramide at 0-0.5, 0.5-2, 2-6, and 6-24 h after tracheal extubation, as well as VAS score for patient satisfaction regarding nausea and vomiting (see attachment, “Study on the Effectiveness of Olanzapine for Preventing Postoperative Nausea and Vomiting” questionnaire).

### 4.2 Study design

Double-blinded, placebo-controlled, parallel-group multicenter study.

### 4.3 Randomization and blinding

#### (a) Randomization

Before beginning participant, an overall assignment manager (designated individual not part of the study group) at Keio university will create computer-generated assignment tables for each center and provide them for each center-specific assignment manager. After obtaining a consent, the investigator shall ask the regulatory manager at each site to register the participant. The regulatory manager shall confirm the eligibility of participant, register the participant and give a participant identification code according to a case registration form. The participant identification code is a unique code for participant determined by each institution. The assignment table randomly divides participant identification code into blocks a 1:1 ratio of olanzapine placebo.

#### (b) Blinding

At each site, an unblinded pharmacist uninvolved in this study will follow assignment table to pack olanzapine (5 mg) and lactose (5 mg) in separate opaque capsules (Kobayashi Capsule Co., Ltd.), accompanied by the assignment manager to minimize errors. Packages will be only labeled with participant identification code from assignment table. The regulatory manager will prescribe the research medication the day before surgery. The Department of Pharmacy will store the packaged capsules and deliver them to the hospital ward upon prescription. On the day of surgery, nurses in charge of study participants will orally administer the appropriate capsule. Thus, only assignment managers and pharmacists who prepare the study medication will be aware of the group assignments. The participants, regulatory managers, anesthesiologists, gynecologists, nurses, and researchers who collect data will be unaware of the group assignments.

### 4.4 Investigational agent information

Name: Generic name: Olanzapine

Trade name: Zyprexa granules

Summary: In this study, subjects will be orally administered olanzapine (5 mg) or placebo in double blind-test capsules once, 2 h before scheduled initiation of anesthesia.

### 4.5 Study period

From the approval date (approval notice is issued) to March 31, 2019 This study duration is projected to include sufficient time (approximately 12–18 months) for obtaining the target sample size (220 subjects at all centers) and to prepare publications of the results, including revisions upon journal submission (approximately 6 months).

### 4.6 Suspension of or withdrawal from the trial

· Trial suspension or withdrawal criteria for <u>individual subjects</u> The patient withdraws her consent to participate in the study.
· Trial suspension or withdrawal criteria for <u>part or all of the trial</u> If serious complications occur that for which a causal relationship with olanzapine cannot be ruled out, the investigator will examine whether it is necessary to suspend or discontinue part or all of the trial.

### 4.7 Management of the investigational agent

Please see section 4.3(b).

### 4.8 Protection of subjects’ privacy

· Anonymization Linkable anonymizing will be performed.
· Creation or non-creation of linking tables (linkable and unlinkable anonymizing) Linking tables will be created.
· Storage of subject list and linking table (containing subject ID numbers and personal information) The personal information manager at each center will store the subject list and linking table in a locked cabinet.
· Required procedure for code breaks (unsealing of the assignment table) due to emergencies The Trial Office will contact investigators at each center, who will then directly contact the assignment manager and personal information manager to perform a code break and quickly identify individual subjects.

### 4.9 Raw data

Raw data is defined as the following information in anesthesia records^1^, medical records^2^, and questionniares^3^.

#### 4.9.1 Patient information

Date of consent^2^, age^1^, height^1^, body weight^1^, American Society of Anesthesiologists Physical Status score^1^, smoking behavior within the past year^1^, history of motion sickness^1^, history of PONV^1^

#### 4.9.2 Surgery, anesthesia and postoperative analgesia information

Date of surgery^1^; diagnosis^1 or 2^; type of surgery^1 or 2^; duration of surgery^1^; duration of anesthesia^1^; dosage of intraoperative fentanyl^1^; dosage of intraoperative remifentanil^1^; usenon-use of peripheral nerve block^1^; time from administration of olanzapine (or placebo) to induction of anesthesia^1 and 2^; intraoperative fluid infusion volume^1^; intraoperative blood transfusion volume^1^; intraoperative blood loss^1^; dosage of intraoperative ephedrine^1^; dosage of intraoperative nicardipine^1^; dosage of intraoperative atropine^1^; dosage of fentanyl from extubation to ward return ^1 and 2^; and dosage of analgesics at 0−0.5, 0.5−2, 2−6, and 6−24 h after extubation^2^

#### 4.9.3 Efficacy indicator information^3^

Presence or absence of PONV at 0-0.5, 0.5-2, 2-6, and 6-24 h after extubation; worst nausea severity experienced at 0−0.5, 0.5−2, 2−6, and 6−24 h after extubation; use and dosage of rescue metoclopramide at 0−0.5, 0.5−2, 2−6, and 6−24 h after extubation; results of patient satisfaction survey for postoperative nausea conducted in the morning on the second postoperative day.

#### 4.9.4 Safety indicator information

Safety indicators comprise the following items up to 48 h after extubation. (This time period was selected because olanzapine has a half-life in blood of approximately 30 h.) Pronounced preoperative drowsiness resulting in difficulty walking to the operating room^2^; hypertension or hypotension requiring treatment^1,2^; bradycardia or tachycardia requiring treatment^1,2^; arrhythmia not observed prior to surgery^1,2^; postoperative agitation^2^; no pre-surgery dry mouth^2^; headache^2^; hepatic dysfunction (abnormalities in AST, ALT, γGTP, or ALP)^2^; no pre-surgery metabolic abnormalities (abnormalities in TG, TC, or blood glucose level)^2^; extrapyramidal symptoms^2^; convulsions^2^.

## 5. Inclusion, exclusion, and withdrawal criteria

### 5.1 Inclusion criteria

See section 2.6.

### 5.2 Exclusion criteria

Patients with diabetes, who have used antiemetics within 24 h pre-surgery, or are currently using olanzapine; patients ≥65 years; women who are pregnant, parturient, or currently nursing.

### 5.3 Withdrawal criteria

Oral administration of olanzapine or placebo occurs only once, prior to surgery. If the subject withdraws consent at any point from the time they provide consent to the start of therapy, she is withdrawn from trial participation and never receives oral administration. Data are not collected, follow-up is not performed, and that subject is not included in statistical analysis. Subject substitution (additional enrollment) is not performed. If, after providing consent, the subject fails to meet eligibility criteria before oral administration, the situation is explained and subject comprehension is ascertained. Investigational therapy (oral administration) is then discontinued, data are not collected, follow-up is not performed, and the individual is excluded from statistical analysis. Subject substitution (additional enrollment) is performed.

## 6. Investigational therapy

### 6.1 Set-up

Subjects will be orally administered olanzapine (5 mg) or lactose as a placebo in double blind-test capsules once, 2 h before scheduled anesthesia initiation.

### 6.2 Concomitant therapy

#### 6.2.1 Therapies permitted before or during clinical trial

Concomitant therapy before clinical trial implementation is not regulated. In the event of PONV, an antiemetic (10 mg metoclopramide) will be administered intravenously the patient’s request. For postoperative pain management, intravenous fentanyl can be administered upon patient request before they return to the ward.

#### 6.2.2 Therapies prohibited before and during clinical trial

No therapies are prohibited for study participants prior to implementing the clinical trial except using antiemetics within 24 h before surgery. Upon induction of anesthesia, all patients must be administered intravenous dexamethasone (6.6 mg) for PONV prophylaxis. Prophylactic intraoperative administration of any other antiemetic is prohibited. Anesthesia must be maintained with sevoflurane. Total intravenous anesthesia and nitrous oxide use are prohibited. Concomitant epidural anesthesia and spinal anesthesia are prohibited. Also prohibited is opioid administration for postoperative analgesia after returning to the ward.

### 6.3 Compliance

Subject drug adherence is not applicable because they will be administered the investigational agent only once. Compliance with protocol will be confirmed during monitoring, conducted at a time stipulated in advance.

## 7. Efficacy assessment

### 7.1 Efficacy indicators

See section 4.1.

### 7.2 Methods for assessing, recording, and analyzing efficacy assessment indicators

#### 7.2.1 Definition

Significant nausea (subjective desire to vomit without expulsive muscular movement) is defined as any score ≥40. Vomiting is the involuntary, forceful expulsion of stomach contents. Vomiting also includes retching, the labored, spasmodic, rhythmic contractions of respiratory muscles without expulsion of gastric contents. A patient is classified as having PONV if she experiences any nausea or vomiting or if she uses rescue antiemetics after tracheal extubation.

#### 7.2.2 Recording information in questionnaires

A ward nurse will visit patients at least 0.5, 2, 6, and 24 h post-extubation and record the following items from the Multicenter Study on the Effectiveness of Olanzapine for Preventing Postoperative Nausea and Vomiting questionnaire (see attached document): 1) PONV presence or absence and use or non-use of antiemetics at 0−0.5, 0.5−2, 2−6, and 6−24 h post-extubation; and 2) worst nausea severity experienced at 0−0.5, 0.5−2, 2−6, and 6−24 h post-extubation. Besides asking patients, antiemetics use is also checked via patient medical records. In the morning on the second postoperative day, a nurse will visit patients again to complete the questionnaire on patient satisfaction related to postoperative nausea.

#### 7.2.3 Entries in case report form

At each center, an investigator blinded to assignments will complete a case report for each patient based on their anesthesia records, questionnaires, and medical records.

#### 7.2.4 Handling of questionnaires and case report forms

The regulatory manager will assemble completed case reports and send them to the Trial Office once a week. Each patient’s questionnaire and case report will be filed as a set.

## 8. Safety assessment

### 8.1 Safety assessment indicators

Safety assessment indicators comprise the presence or absence of pronounced preoperative drowsiness, resulting in difficulty walking to the operating room, and the presence or absence of the following phenomena from start of investigational therapy to 48 h after extubation.

∘ Intraoperative hypertension, hypotension, tachycardia, and bradycardia requiring treatment
∘ Postoperative agitation
∘ Postoperative sedation
∘ Postoperative dry mouth
∘ Postoperative headache
∘ Postoperative rise or fall in blood pressure requiring treatment
∘ Arrhythmia not observed pre-surgery
∘ Hepatic dysfunction (abnormalities in AST, ALT, γGTP, or ALP) not observed pre-surgery
∘ Metabolic abnormalities (abnormalities in TG, TC, or blood glucose level) not observed pre-surgery
∘ Extrapyramidal symptoms
∘ Convulsions

### 8.2 Methods for assessing, recording, and analyzing safety assessment indicators

Presence or absence of the items listed in 8.1 will be entered in the case report. The subsequent analysis will determine incidence of each item.

### 8.3 Adverse events

#### 8.3.1 Definition of adverse event (AE)

Please see section 8.1.

#### 8.3.2 Definition of serious adverse event (SAE)

An SAE is defined as any AE (8.3.1) occurring up to 48 h post-extubation meeting any of the following criteria.

∘ AE resulting in death
∘ Life-threatening AE
∘ AE requiring hospitalization or extension of current hospitalization for treatment
∘ AE resulting in permanent or severe injury or dysfunction
∘ AE causing congenital abnormalities

#### 8.3.3 Procedure for collecting, recording, and reporting AE or SAE

##### A) Events that must be reported

All SAE, excluding those for which a causal relationship with olanzapine can be ruled out.

##### B) Reporting method

Reports will be made in accordance with the procedure for reporting SAE in clinical studies involving invasiveness (including advanced medical care).

##### C) Report destination

Reports will be sent to the Trial Office. The Trial Office will report to investigators and study center directors at all centers.

## 9. Statistical analysis

(See attached statistical analysis plan for more detailed information.)

### 9.1 Analysis method

A statistician uninvolved in the study or data collection will analyze results based on information recorded in case reports.

### 9.2 Sample size

220 patients

If PONV incidence within 24 h of extubation in the control group is 60%, then olanzapine reduction of PONV incidence to 40% is considered clinically effective. If α = 5% and β = 20%, the sample size is 97 subjects per group. Assuming a dropout rate of approximately 10%, each group requires 110 subjects.

Target sample sizes for each center

· Keio University Hospital: 70
· Showa University Hospital: 60
· Jikei University Hospital: 40
· University Hospital, Kyoto Prefectural University of Medicine: 30
· Nagoya City University Hospital: 20

### 9.3 Level of statistical significance

Significance is set at p < 0.05 in two-tailed testing.

### 9.4 Handling of trial data

In the event of missing values, the data in question will be excluded from analysis.

### 9.5 Deviation from statistical analysis protocol

Details regarding excluded data will be described in detail when publishing the report.

### 9.6 Analysis set

· Subject selection and definition of evaluable cases All subjects who provide consent, are orally administered olanzapine or placebo, and undergo surgery.

## 10. Direct viewing of raw data and raw materials

Raw data and materials will be submitted for direct viewing upon monitoring and auditing related to the clinical study, review by ethics committees, as well as inspection by the Ministry of Health, Labour and Welfare.

## 11. Monitoring

Quality control monitoring will be conducted using the following methods.

∘ Monitor: study group per study center
∘ Timing and frequency: monitoring performed for three cases per center after thestart of the study, followed by monitoring of three cases every 6 months from approval date. Inclusion criteria, exclusion criteria, and primary outcome will be determined for each subject.
∘ Monitoring methods: see attached monitoring procedure manual.
∘ Report destination: Trial Office

## 12. Ethics

### 12.1 Informed consent

This study will explained both in writing and orally to patients either upon an outpatient visit for an anesthesia explanation, or during an anesthesia explanation in a gynecology ward. Patients will then be asked to participate in the study.

### 12.2 Reporting to the ethical committee

· Annual reports Study progress will be reported to the ethical committee at each center once a year after approval is obtained.
· Reports of suspension/withdrawal/conclusion Directors of study centers will be notified of suspensions, withdrawals, or conclusions.

### 12.3 Publication of information related to the study

· Clinical trial enrollment
  ∘ The study will be registered with and publicized on UMIN before the study begins.
  ∘ Information related to the present study will be published on the home page of the Department of Anesthesiology, Keio University School of Medicine.

### 12.4 Corresponding investigator for inquiries from subjects and related individuals

· Name: Yuya Masuda
· Affiliation: Dept. of Anesthesiology, Keio University School of Medicine
· Position: Assistant Professor
· Contact information: 35 Shinanomachi, Shinjuku-ku, Tokyo 160-5852, Japan
· TEL: +81-3-5363-3107
· FAX: +81-3-3356-8439
· E-mail address

## 13. Handling of data and samples, and preservation of records

Questionnaires, case reports, linking tables, and assignment tables will be stored in a locked cabinet at each center for the longest duration of the following situations: at least 5 years following the date of the study conclusion report, or 3 years following the date of the final report of study results. Subsequently, these documents will be shredded.

## 14. Financial burden on subjects and measures other than insurance

· Financial burden imposed on subjects by trial participation None
· Issuance of reimbursement (compensation) to subjects for participation None
· Measures other than insurance for health damage compensation Covered by clinical trial insurance

## 15. Arrangement for publication of trial results

The results obtained in this study will be presented at conferences and published in papers; individual subject identifying information will not be revealed.

## 16. Addenda

The olanzapine dose generally used to prevent CINV is 10 mg, a concentration considered safe (Rudolph L, et al., N Engl J Med 2016;375:134-142). This dose was also used in a previous study that examined olanzapine effectiveness for preventing PONV (Ibrahim M, et al. Egypt J Anaesth 2013;29:89-95). In addition, 10 mg does not exceed the amount recommended on the olanzapine package insert. For these reasons, we initially set the olanzapine dose at 10 mg. In addition, we originally administered the drug 4 h before anesthesia induction because T_max_ following oral administration of olanzapine is approximately 5 h. However, following the start of the study, three patients demonstrated pronounced preoperative drowsiness transport to the operating room using a wheelchair or stretcher. This drowsiness was suspected to be associated with olanzapine. We also noted that many studies on olanzapine effects used non-Japanese data, suggesting that our Japanese subjects may require a lower dose. Therefore, we reduced the olanzapine to 5 mg, the minimum dosage on the package insert. To maximize blood concentration of olanzapine at the end of anesthesia, we changed the timing of administration to 2 h before anesthesia induction.

**Multicenter Study on the Effectiveness of Olanzapine for Preventing Postoperative Nausea and Vomiting: Statistical Analysis Plan**

## 1. Introduction

This statistical analysis plan describes in detail the statistical methods that are outlined in the version 3.0 protocol of “Multicenter Study on the Effectiveness of Olanzapine for Preventing Postoperative Nausea and Vomiting: A Clinical Trial Protocol,” 9. Statistical analysis.”

## 2. Aim of the study

The purpose of this study is to examine whether oral olanzapine reduces postoperative nausea and vomiting (PONV).

## 3. Definition

Nausea was defined as a subjective feeling of a desire to vomit without the presence of expulsive muscular movements. Vomiting was defined as the involuntary, forceful expulsion of the contents of stomach. Retching was defined as the labored, spasmodic, rhythmic contractions of the respiratory muscles without the expulsion of gastric contents and was included in vomiting. A patient was considered to have PONV if she experienced any episodes of nausea or vomiting, or if she used rescue antiemetics after tracheal extubation.

## 4. Study design and participants

This study is a multi-center, randomized, double-blinded, parallel-group, placebo-controlled clinical trial. Patients aged 20–65 years who underwent elective laparoscopic gynecological surgery expected to last at least 1 h for a benign indication who had ≥2 risk factors according to the simplified Apfel score were eligible for inclusion in this study. Patients were excluded if they were diagnosed with diabetes, had taken preoperative olanzapine outside the study protocol, had contraindications to any of the study drugs, had taken emetogenic or antiemetic drugs within 24 h of surgery, had undergone emergency surgery, or were pregnant or lactating.

## 5. Outcomes

### 5.1 Primary outcome

The primary outcome measure is the proportion of study patients who have PONV within a 24-h postoperative period.

### 5.2 Secondary outcome

1. Time to first PONV
2. Incidence of PONV at 0–0.5, 0.5–2, 2–6, and 6–24 h after surgery
3. Incidence of vomiting at 0–0.5, 0.5–2, 2–6, and 6–24 h after surgery
4. nausea VAS score at 0–0.5, 0.5–2, 2–6, and 6–24 h after surgery
5. Incidence of severe nausea (VAS score for nausea ≥40) at 0–0.5, 0.5–2, 2–6, and 6– 24 h after surgery
6. Use and dose of rescue metoclopramide at 0–0.5, 0.5–2, 2–6, and 6–24 h after surgery
7. The VAS score for patient satisfaction regarding nausea

### 5.3 Adverse events (AE)s

Incidences of

1. Pronounced preoperative drowsiness resulting in difficulty walking to the operating room
2. Intraoperative hypotension requiring treatment
3. Intraoperative hypertension requiring treatment
4. Intraoperative arrhythmia that was not preexisting
5. Postoperative agitation (up to 48 h after extubation)
6. Postoperative sedation (up to 48 h after extubation)
7. Postoperative dry mouth (up to 48 h after extubation)
8. Postoperative headache (up to 48 h after extubation)
9. Postoperative extrapyramidal symptoms (up to 48 h after extubation)
10. Postoperative seizures (up to 48 h after extubation)
11. Postoperative abnormal laboratory values specified by the protocol (alanine aminotransferase, aspartate aminotransferase, gamma-glutamyl transpeptidase, alkaline phosphatase, triglyceride, total cholesterol, and blood glucose) not observed pre-surgery (up to 48 h after extubation)

### 5.4 Serious AEs

Incidences of

1. AE resulting in death
2. Life-threatening AE
3. AE requiring hospitalization or extension of current hospitalization for treatment
4. AE resulting in permanent or severe injury or dysfunction
5. AE causing congenital abnormalities

*SAE for which a causal relationship with olanzapine can be ruled out shall be excluded from analysis.

## 6. Statistical methods and reporting

### 6.1 Sample size

A power analysis was performed with power = 80% and α = .05. Given our retrospective observation that 61% of patients who received general anesthesia developed PONV within 24 h of surgery, we assumed that the incidence of PONV within 24 h of surgery would be approximately 60% for the control group. We considered that a 20% reduction in the absolute risk of PONV would have clinical significance. The power analysis showed that 97 patients were needed in each group. Allowing for possible dropouts, we will allocate 110 patients to each group.

### 6.2 Level of significance

For all analyses used for assessment of primary and secondary endpoints, a p-value of less than 0.05 (two-tailed) will be considered statistically significant.

### 6.3 Study analysis set

The modified intention to treat (m-ITT) population, defined as all randomized participants who received either olanzapine or placebo and who underwent the planned surgical procedure, was used for the primary dataset for the analysis.

### 6.4 Items and methods for analysis

#### 6.4.1 Baseline characteristics of the participants

The baseline characteristics of the participants will be summarized by standard descriptive statistics (e.g., means [standard deviation] or median [interquartile range] for continuous variables, and percentage for categorical variables). No statistical comparison between the two study groups will be made since there is no hypothesis being tested at baseline. The baseline variables that are relevant to PONV will be included as a covariate in the multivariable analysis to account for any imbalance that may have occurred by chance between the treatment groups.

The following demographic data will be collected and be presented:

- age
- height
- body weight
- American Society of Anesthesiologists Physical Status score
- smoking behavior within the past year
- history of motion sickness
- history of PONV
- number of risk factors for PONV

#### 6.4.2 Perioperative variables of the participants

The perioperative variables of the study patients will be summarized by standard descriptive statistics (e.g., means [standard deviation] or median [interquartile range] for continuous variables). The standardized difference between the two study groups will be presented. The perioperative variables that are relevant to PONV will be included as covariates.

The following variables will be collected and be presented:

- duration of surgery
- duration of anesthesia,
- dosage of intraoperative fentanyl
- dosage of intraoperative remifentanil
- time from administration of olanzapine (or placebo) to induction of anesthesia
- intraoperative fluid infusion volume
- intraoperative blood transfusion volume
- intraoperative blood loss
- dosage of intraoperative ephedrine
- dosage of intraoperative nicardipine
- dosage of intraoperative atropine
- dosage of fentanyl from extubation to ward return

#### 6.4.3 Analysis principles

All analyses will be performed according to the intention-to-treat (ITT) principle using the m-ITT population. The data will be analyzed according to the trial arm to which the participant was randomized. All tests will be two-sided, and results will be considered as statistically significant using the 5% level. There will be no imputation for missing data. There will be no adjustment for multiple testing. p-values will be reported to four decimal places with p-values less than 0.0001 as <0.0001. The statistical software R with appropriate add-on packages will be used for analyses.

#### 6.4.4 Analysis of primary outcome measure

The primary outcome is the incidence of PONV within 24 h after extubation. The Cochran-Mantel-Haenzel test will be employed for stratified analyses to take potential differences across hospitals into account and to assess the homogeneity of the treatment effect. Using the Cochran-Mantel-Haenzel test, the adjusted odds and risk ratios with 95% confidential intervals will be calculated. The heterogeneity across different hospitals will be examined by χ2 test. For sensitivity analysis of primary outcome measure, multivariate logistic regression analysis will be conducted. The covariates for multivariate adjustment will be chosen from variables that have clinical relevance to PONV. The step-wise procedure based on minimization of the AIC (Akaike Information Criterion) will be applied for the model selection.

#### 6.4.5 Analysis of secondary outcome measures

Statistical analysis methods for each secondary outcome will be as follows:

##### 1) Time to first PONV

Time to event analysis will be carried out with the Kaplan-Meier method, using a log-rank test for difference of curve pairs. If appropriate, adjustment for multiple confounders may be conducted using the Cox proportional hazard regression model with a backward stepwise procedure. The treatment effect of OLZ will be quantified by hazard ratio with 95% confidence interval.

##### 2) Changes in PONV risk within 24 hours after surgery

For each patient, a binary sequence of PONV in the four consecutive observation periods (0–0.5, 0.5–2, 2–6, and 6–24 h after surgery) will be constructed. We expect an extremely low missing data rate, as the study periods are during the immediate post-operative phase and all patients will be under close observation.

The goal of the analysis will be to determine subject-specific changes in the risk of PONV over the course of the post-operative period and the influence of OLZ on changes in a woman’s risk of PONV. The following mixed effects logistic regression model will be constructed and parameters will be estimated with the maximal likelihood method. The model posits natural heterogeneity in a woman’s underlying risk of PONV that persists throughout the study periods.

**Generalized linear mixed effects model to account for the changes in PONV risk during the post-operative period**

1. conditional mean response: logistic linear model with random intercept

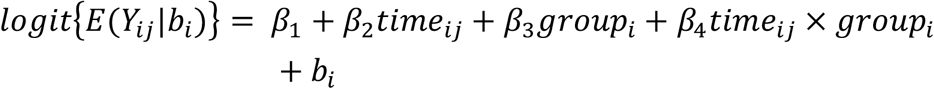

Y_ij_ : Y_ij_=1 if the i^th^ patient experienced PONV in the j^th^ occasion, otherwise Y_ij_=0

i : patient

j : measurement occasion (j=0∼3)

time_ij_ : i^th^ patient, j^th^ measurement occasion

group_i_ : treatment group of i^th^ patient

(group_i_=1 if i^th^ patient is OLZ, otherwise group_i_=0)

b_i_: random intercept, b_i_ ∼ N (0, σ_b_^2^)

2. variance function

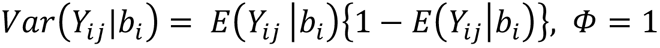

Φ: scale parameter

##### 3) Changes in vomiting risk within 24 hours after surgery

For each patient, a binary sequence of vomiting in the four consecutive study periods (0-0.5, 0.5-2, 2-6, and 6-24 hours after surgery) will be constructed. In order to determine subject-specific changes in the risk of vomiting over the course of the post-operative period and the influence of OLZ on changes in a woman’s risk of vomiting, the mixed effects logistic regression model with a random intercept will be constructed to examine treatment group by time interaction. The parameters will be estimated with maximal likelihood method.

**Generalized linear mixed effects model for accounting for the changes in vomiting risk during post-operative period**

1. conditional mean response: logistic linear model with random intercept

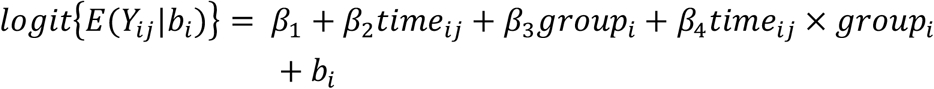

Y_ij_ : Y_ij_=1 if the i^th^ patient experienced vomiting in the j^th^ occasion, otherwise Y_ij_=0

i : patient

j : measurement occasion (j=0∼3)

time_ij_ : i^th^ patient, j^th^ measurement occasion

group_i_ : treatment group of i^th^ patient

(group_i_=1 if i^th^ patient is OLZ, otherwise group_i_=0)

b_i_: random intercept, b_i_ ∼ N (0, σ_b_^2^)

2. variance function

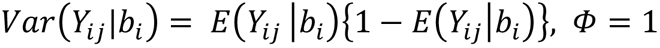

Φ: scale parameter

##### 4) Changes in severe nausea risk within 24 h after surgery

In this study, severe nausea is defined as a VAS score for nausea greater than or equal to 40. For each patient, a binary sequence of severe nausea in the four consecutive study periods (0–0.5, 0.5–2, 2–6, and 6–24 h after surgery) will be constructed.

We will evaluate the effect of OLZ on the trajectory of severe nausea using a mixed effects logistic regression model, with individual patient as a random effect. The parameters will be estimated with the maximal likelihood method.

**Generalized linear mixed effects model to account for the changes in severe nausea risk during the post-operative period**

1. conditional mean response: logistic linear model with random intercept

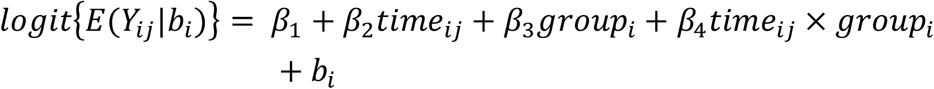

Y_ij_ : Y_ij_=1 if the i^th^ patient experienced severe nausea in the j^th^ occasion, otherwise Y_ij_=0

i : patient

j : measurement occasion (j=0∼3)

time_ij_ : i^th^ patient, j^th^ measurement occasion

group_i_ : treatment group of i^th^ patient

(group_i_=1 if i^th^ patient is OLZ, otherwise group_i_=0)

b_i_: random intercept, b_i_ ∼ N (0, σ_b_^2^)

2. variance function

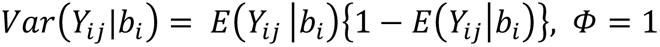

Φ: scale parameter

##### 5) Longitudinal analysis of rescue metoclopramide administration within 24 h after surgery

For each patient, a binary sequence of metoclopramide administration in the four consecutive study periods (0–0.5, 0.5–2, 2–6, and 6–24 h after surgery) will be constructed.

A mixed effects logistic regression model will be used to determine longitudinal trends in rescue metoclopramide administration and the influence of OLZ on the PONV rescue treatment. The parameters will be estimated with the maximal likelihood method.

**Generalized linear mixed effects model to account for the changes in metoclopramide administration during the post-operative period**

1. conditional mean response: logistic linear model with random intercept

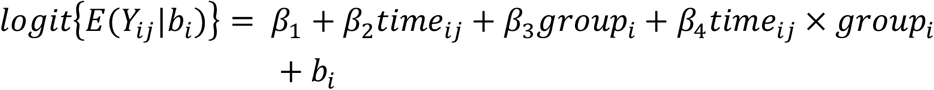

Y_ij_ : Y_ij_=1 if the i^th^ patient having metoclopramide in the j^th^ occasion, otherwise Y_ij_=0

i : patient

j : measurement occasion (j=0∼3)

time_ij_ : i^th^ patient, j^th^ measurement occasion

group_i_ : treatment group of i^th^ patient

(group_i_=1 if i^th^ patient is OLZ, otherwise group_i_=0)

b_i_: random intercept, b_i_ ∼ N (0, σ_b_^2^)

2. variance function

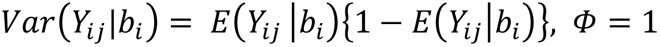

Φ: scale parameter

##### 6) Changes in VAS score for nausea within 24 h after surgery

The VAS score for nausea will be considered a continuous variable. To account for the difference of VAS trajectory in two treatment groups, a general linear mixed effects model will be constructed and will include the randomized study group, time points, interaction between the study group, and time points as fixed effects, with individual as a random effect. The parameters will be estimated with the restricted maximal likelihood method.

**Generalized linear mixed effects model to account for the changes in the severity of nausea (VAS score) during the post-operative period**

1. conditional mean response: general linear model with random slope and intercept

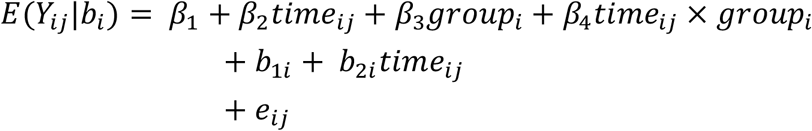

*E*(*Y*_*i,j*_*b*_*i*_): conditional mean VAS value of i^th^ patient in the j^th^ occasion

*i* : patient

*j* : measurement occasion (j=0 3)

*time*_*ij*_ : i^th^ patient, j^th^ measurement occasion

*group*_*i*_ : treatment group of i^th^ patient

(group_i_=1 if i^th^ patient is OLZ, otherwise group_i_=0)

*b*_*1i*_: *random intercept, b*_*1i*_ *∼ N (0*, _*b1*_^*2*^*) b*_*2i*_: *random slope, b*_*2i*_ *∼ N (0*, _*b2*_^*2*^)

*e*_*ij*_: *random error*

2. variance function

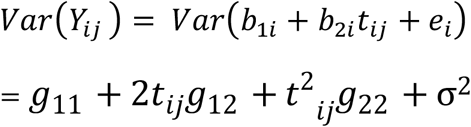

*Var*(*b*_1*i*_) = *g*_11_

*Var*(*b*_2*i*_) = *g*_22_

*cov*(*b*_1*i*,_b_2*i*_) = *g*_12_

assume *cov*(*e*_*ij*,_e_*ik*_) = 0 *and var* (*e*_*ij*_*) =* σ^2^

#### 6.4.5 Adverse events

The incidence of AEs and SAEs will be analyzed with the m-ITT population and summarized by treatment group.

## 7. Interim analysis

No interim analysis is planned.

## Data Availability

Data available: No.
Explanation for why data not available: The informed consent form signed by the patients did not address an individual data sharing statement.

## List of abbreviations

ALP: Alkaline phosphatase
ALT: Alanine aminotransferase
AST: Aspartate aminotransferase
γGTP: Gamma-glutamyl transpeptidase
CINV: Chemotherapyinduced nausea and vomiting
Dept: Department
PONV: Postoperative nausea and vomiting
TG: Triglyceride
TC: Total cholesterol
UNMIN: University Hospital Medical Information Network
VAS: Visual analogue scale

